# Antigen-based testing but not real-time PCR correlates with SARS-CoV-2 virus culture

**DOI:** 10.1101/2020.10.02.20205708

**Authors:** Andrew Pekosz, Charles K. Cooper, Valentin Parvu, Maggie Li, Jeffrey C. Andrews, Yukari C. Manabe, Salma Kodsi, Jeffry Leitch, Devin S. Gary, Celine Roger-Dalbert

**Affiliations:** W. Harry Feinstone Department of Molecular Microbiology and Immunology, Johns Hopkins Bloomberg School of Public Health; Department of Emergency Medicine; Becton, Dickinson and Company, BD Life Sciences – Integrated Diagnostic Solutions, 7 Loveton Circle, Sparks, MD, USA; Department of Medicine, Johns Hopkins University School of Medicine, Baltimore, Maryland

**Keywords:** SARS-CoV-2, Antigen testing, SARS-CoV-2 contagious, viral load, live culture, RT-PCR testing

## Abstract

Individuals can test positive for SARS-CoV-2 by real-time polymerase chain reaction (RT-PCR) after no longer being infectious.^1-8^ Positive SARS-CoV-2 antigen-based testing exhibits a temporal pattern that corresponds with active, replicating virus and could therefore be a more accurate predictor of an individual’s potential to transmit SARS-CoV-2.^2,3,9^ Using the BD Veritor System for Rapid Detection of SARS-CoV-2 later flow antigen detection test, we demonstrate a higher concordance of antigen-positive test results with the presence of cultured, infectious virus when compared to RT-PCR. When compared to infectious virus isolation, the sensitivity of antigen-based testing is similar to RT-PCR. The correlation between SARS-CoV-2 antigen and SARS-CoV-2 culture represents a significant advancement in determining the risk for potential transmissibility beyond that which can be achieved by detection of SARS-CoV-2 genomic RNA. Coupled with a rapid time-to-result, low cost, and scalability, antigen-based testing should facilitate effective implementation of testing and public health interventions that will better contain COVID-19.

## INTRODUCTION

The SARS-CoV-2 causes COVID-19 and is spread from human-to-human primarily through airborne transmission.^10^ The mean incubation time, or presymptomatic period, for SARS-CoV-2 is approximately 5.8 days (95% CI 5.0-6.7),^11,12^ and the period of transmission (the total time during which a patient is contagious) begins around one to three days prior to symptom onset, with a subsequent reduction in contagiousness occurring 7-10 days following symptom onset.^8,13,14^ Recent work in a golden hamster SARS-CoV-2 model demonstrated that although the presence of genomic RNA in nasal washes extends to 14 days post-inoculation, the detection of infectious virus and the communicable period both end well before 14 days.^15^ In addition, four previous studies, utilizing culture-based virus detection from human specimens, demonstrated an absence of infectious isolates from most specimens taken eight days after symptom onset, despite measurable viral RNA loads using RT-PCR.^1,3,5,7^

Several SARS-CoV-2 antigen-based tests, which work via a lateral flow immunoassay mechanism, have recently received Emergency Use Authorization (EUA) from the Food and Drug Administration.^16-19^ Several lines of indirect evidence suggest that antigen-based testing may align better with culture-based test results compared to RT-PCR. For example, higher RT-PCR Ct values from specimens are observed when individuals are negative by antigen testing or culture-based testing compared to those from individual that are antigen test ^9^ or culture–based test positive.^6^ In addition, current EUA SARS-CoV-2 antigen tests have optimal performance profiles^16-19^ at time points that overlap with the temporal expression profile of SARS-CoV-2 sub-genomic RNA (a marker for active, replicating virus).^3^ Despite the recognition that point-of-care or other testing modalities might be more effective at discerning contagious from non-contagious individuals,^2^ no study has directly compared antigen-based testing with RT-PCR in the same study using a reference method for infectiousness.

The objective of this study was to determine whether SARS-CoV-2 antigen testing differentiates SARS-CoV-2-contagious individuals (e.g., those still shedding infectious virus) from non-contagious individuals compared to RT-PCR methodology. To address this, we utilized Quidel Lyra^**®**^ SARS-CoV-2 Assay (“RT-PCR assay”) positive and negative specimens obtained from a diverse set of collection sites across the USA. The RT-PCR assay and the BD Veritor™ System for Rapid Detection of SARS-CoV-2 (“antigen test”) were compared to SARS-CoV-2 TMPRSS2 culture (a sensitive virus culture test utilizing the VeroE6TMPRSS2 cell line), which served as the reference method for determining infectiousness.

## RESULTS

The 38 RT-PCR positive specimens were tested for the presence of SARS-CoV-2 using infection of VeroE6TMPRSS2 cell cultures (SARS-CoV-2 TMPRSS2 culture). Overall, 28 RT-PCR-positive specimens were also positive by SARS-CoV-2 TMPRSS2 culture and 10 of 38 RT-PCR-positive specimens were negative by SARS-CoV-2 TMPRSS2 culture (Figure 1A). SARS-CoV-2 TMPRSS2 culture-positive specimens had a mean log10 viral RNA copy number of 7.16 compared to 4.01 from specimens that were SARS-CoV-2 TMPRSS2 culture-negative (p-value <0.001; two-sample t-test, 2-tailed analysis). Further stratification by results from the antigen test showed that 27 of 28 RT-PCR-positive/SARS-CoV-2 TMPRSS2 culture-positive specimens were also positive in the antigen test; only two of the ten RT-PCR-positive/ SARS-CoV-2 TMPRSS2 culture-negative specimens were positive by the antigen test.

**Figure 1.**
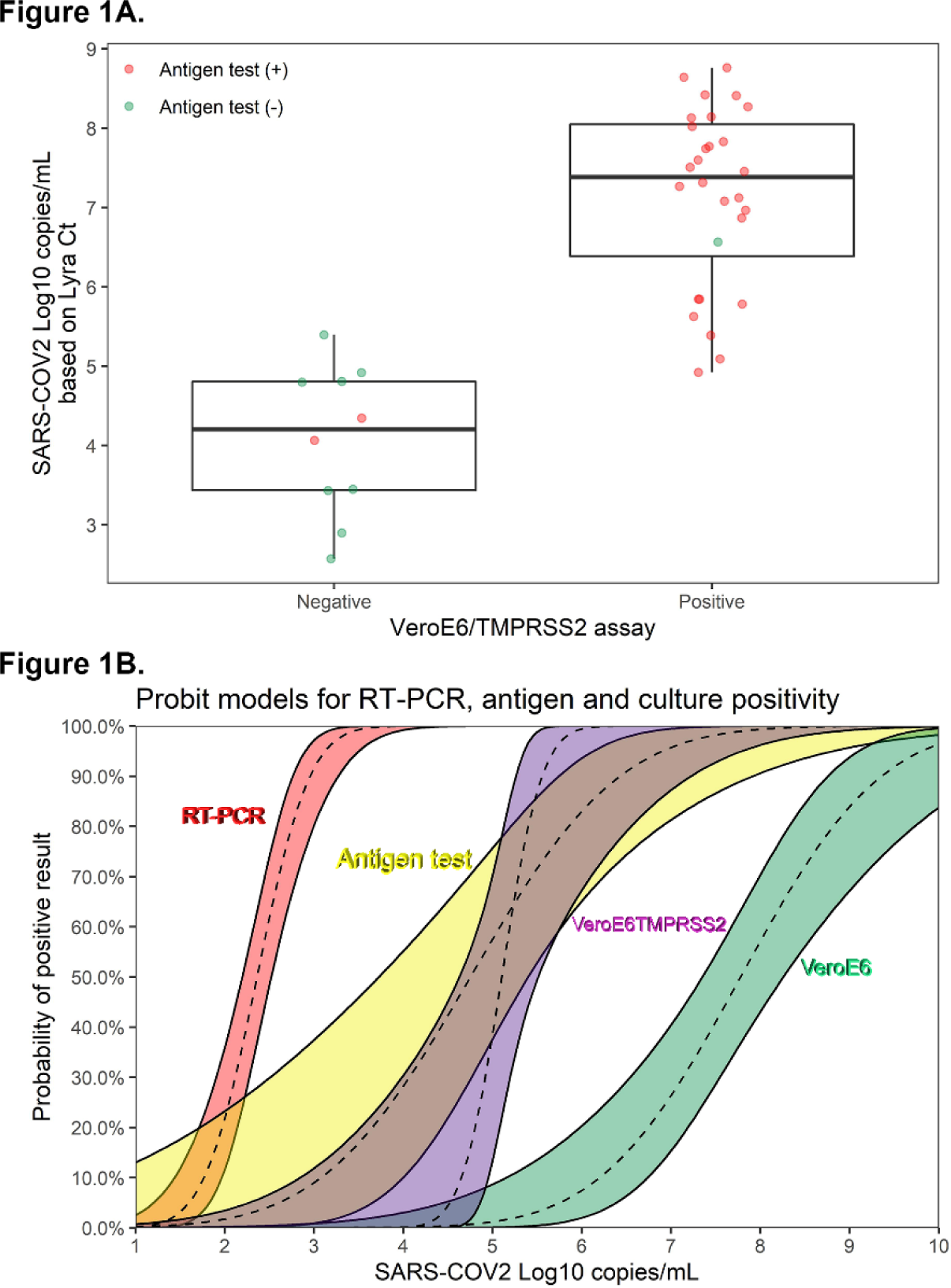
(A) The 38 RT-PCR assay positive specimens are plotted by Log10 copies/mL (y-axis) and are stratified by the SARS-CoV-2 live culture results (negative, n=10; positive, n=28). The median and inter-quartile range values, respectively, for the RT-PCR-positive/SARS-CoV-2 TMPRSS2 culture-negative were 4.21 and 1.37; the median and inter-quartile range values, respectively, for the RT-PCR-positive/SARS-CoV-2 TMPRSS2 culture-negative were 7.39 and 1.66. The mean values for the SARS-CoV-2 TMPRSS2 culture-negative and SARS-CoV-2 TMPRSS2 assay-positive specimen groups were significantly different (4.01 versus 7.16, respectively; p-value <0.001 based on two-sample t-test [2-tailed]). Antigen test positive results are indicated as red data points (n=29) and the antigen test negative results (n=9) are indicated by the green data points. (B) Probit models linking viral load to the probability of positive result of RT-PCR (Lyra), antigen test (Veritor), SARS-CoV-2 TMPRSS2 culture and SARS-CoV-2 VeroE6 culture (refer to Huang et al 2020). Viral load levels at which there is a 5% chance of positive result: 1.6, 2.6, 4.5, and 5.75 log10 cp/ml for RT-PCR, antigen, SARS-CoV-2 TMPRSS2 culture, and SARS-CoV-2 VEroE6 culture, respectively.

Of the 38 RT-PCR-positive results utilized for these analyses, nine were antigen test negative. These nine negative results showed a trend towards longer time from symptom onset compared to the 29 RT-PCR assay-positive/antigen test-positive specimens (4.4 days on average versus 2.9, p-value = 0.108).^9^ Of the nine samples that were RT-PCR-positive/antigen test-negative, the viral RNA copy number was significantly lower than that observed for the 29 RT-PCR-positive/antigen test-positive specimens (mean 4.3 log10 cp/mL versus 7.0 log10 cp/mL, p-value<0.001, Figure S1). Symptom day was not a significant factor in multivariate models, while viral RNA load continued to be significant (p-value = 0.002).

Probit models for percent positivity by viral RNA load corresponding to the RT-PCR assay, antigen test, SARS-CoV-2 TMPRSS2 culture, and SARS-CoV-2 culture with VeroE6 cells (“SARS-CoV-2 VeroE6 culture;” data integrated into the probit model using previous data; see Methods)^6^ are provided in Figure 1B. The SARS-CoV-2 VeroE6 culture yielded a positive result at a rate of 5% for a viral load of 5.75 log10 cp/ml, whereas the SARS-CoV-2 TMPRSS2 culture corresponded to a positive result with a rate of 5% at a viral load of 4.5 log10 cp/mL. At a viral load of 2.6 log10 cp/mL, the antigen test yielded a positive result at a rate of 5%. In a multivariate generalized linear model with viral RNA load and test type, the SARS-CoV-2 TMPRSS2 culture was not significantly different from the antigen test (p-value = 0.953). Both the SARS-CoV-2 TMPRSS2 culture and antigen test were significantly more likely to yield positive results than SARS-CoV-2 VeroE6 culture (p-value<0.001 for both). Unlike the antigen test, the RT-PCR assay showed very little overlap with SARS-CoV-2 TMPRSS2 culture, yielding positive results at much lower viral loads.

As shown in Table 1, the antigen test demonstrated a sensitivity and specificity of 96.4% (95% CI: 82.3, 99.4) and 98.7% (96.1, 99.7), respectively. The RT-PCR assay demonstrated a sensitivity and specificity of 100% (87.7, 100) and 95.5% (91.1, 97.8), respectively. Based on the study prevalence of 11.2%, as determined by SARS-CoV-2 TMPRSS2 culture positivity and a total specimen number of 251 (based on the total, evaluable specimen set utilized for the Veritor EUA study), the positive predictive value (PPV) for the antigen test was 90.0% (76.3, 97.6), while the PPV for the RT-PCR assay was only 73.7% (60.8, 85.3).

**Table 1.** Performance of the antigen test and the RT-PCR assay for detection of SARS-CoV-2 infectivity based on virus culture positive results within 0-7 days from symptom onset

| **Prevalence:** 11.2% |  |  |
| --- | --- | --- |
| **Performance values** | **Antigen test performance** | **RT-PCR assay performance** |
| **Sensitivity** | 96.4 [82.3, 99.4] | 100 [87.7, 100] |
| **Specificity** | 98.7 [96.1, 99.7] | 95.5 [91.1, 97.8] |
| **PPV** | 90.0 [76.3, 97.6] | 73.7 [60.8, 85.3] |
| **NPV** | 99.5 [97.7, 100] | 100 [98.4, 100] |
| **Accuracy** | 98.4 [96.0, 99.4] | 96.0 [92.8, 97.8] |
| **TP** | 27 | 28 |
| **FP** | 3 | 10 |
| **FN** | 1 | 0 |
| **TN<sup>a</sup>** | 220 | 213 |

**Abbreviations:** RT-PCR, real-time polymerase chain reaction; PPV, positive predictive value; NPV, negative predictive value; TP, true positive; FP, false positive; FN, false negative; TN, true negative
<sup>a</sup>includes 176 specimen sets that were RT-PCR and antigen negative, with unavailable culture results.

## DISCUSSION

The results here show similar sensitivity between the SARS-CoV-2 antigen test and the SARS-CoV-2 RT-PCR assay (96.4% and 100%, respectively) over a time range of <8 days post symptom onset. However, the SARS-CoV-2 antigen test had a PPV of 90.0%, whereas the RT-PCR assay showed a PPV of only 73.7%. In addition, the probit model for percent positivity employed in this study showed considerable overlap between the antigen test and the SARS-CoV-2 TMPRSS2 culture, with little overlap between the SARS-CoV-2 TMPRSS2 culture and RT-PCR.

Ct values are inversely correlated with the viral load thresholds corresponding to infectious virus isolation. Because limits of detection vary between RT-PCR assays, however, Ct values reported by specific RT-PCR assays correspond to different viral RNA loads.^6,7,20-25^ Here we utilized the Lyra assay to establish a probit model of percent positivity by viral load, which facilitates a better comparison of these results with previous work. Recent studies involving upper respiratory swab specimens reported no cases of COVID-19 with SARS-CoV-2 viral RNA loads below 4 log10 cp/mL.^1,3,5,6,8,13,26,27^ Other work has shown that specimens with viral RNA loads ≤6 log10 cp/ml have minimal or no culturable SARS-CoV-2 virus.^3,5,28-30^ Here, a low percent positivity (5-10%) was observed for the SARS-CoV-2 VeroE6-culture test below 6 Log10 cp/mL. The SARS-CoV-2 TMPRSS2 culture test, however, showed 90% positivity at 5.6 log10 cp/mL. Although antigen test had a larger distribution of positivity, it overlapped considerably with the SARS-CoV-2 TMPRSS2 culture test and approached 90% positivity at a viral RNA load of 6.4Log10. This is consistent with the WHO target product profile for priority diagnostics, which supports viral RNA load based methodologies and includes an acceptable limit of detection for point-of-care tests of 6Log10 cp/mL.^31^

As with other viruses, RT-PCR-based methodologies may be detecting SARS-CoV-2 RNA even after infectious virus is no longer present;^32-37^ especially at time periods beyond 7 days from symptom onset.^1,3^ For most patients with COVID-19, efforts to isolate live virus from upper respiratory tract specimens have been unsuccessful ≥10 days from symptom onset; it is unlikely that these individuals pose a transmission risk to others.^25^ In addition, there is no evidence to date that persistent or recurrent detection of viral RNA, following recovery from COVID-19, poses a risk of SARS-CoV-2 transmission.^25^

This work highlights a key potential value of decentralized POC antigen-based testing and furthers our understanding of the interpretation of antigen test results. Antigen testing facilitates accurate and rapid detection of infectious individuals who may not require direct medical management (due to mild/non-severe disease), but for whom infection control measures have the potential to interrupt community transmission. While RT-PCR is highly sensitive when compared to SARS-CoV-2 TMPRSS2culture, antigen testing also showed excellent sensitivity (96.4%) coupled with better PPV relative to RT-PCR (90.0 versus 73.7) and rapid time to results.

This study had limitations. It only included specimens from patients within seven days of symptom onset. Several studies have demonstrated an inability to culture SARS-CoV-2 beyond day eight, despite ongoing RT-PCR positivity.^1,3,5^ Serial sampling of COVID-19 patients is needed to determine if there is a propensity to have viral antigen test positive results after a negative result, as can sometimes be seen with RT-PCR tests. Results from this study likely underestimate the difference in specificity between RT-PCR and antigen testing that would be expected in a set that included specimens collected at later times post symptom onset. In this study, while three subjects were antigen test false positives versus SARS-CoV-2 TMPRSS2 culture, as many as ten subjects were RT-PCR false positives versus culture (viral RNA loads ranging from 2.6 to 5.4 log10 copies/mL). Although the sample size was adequate in this study, the confidence intervals in the probit model were too wide to establish a definitive viral load cut-off. To improve the precision associated with the point estimates, either a larger study or a meta-analysis, involving multiple studies, would be required. Also, there are limitations associated with the use of culture positivity or viral RNA load as a surrogate for infectiousness or transmissibility that require further investigation. Finally, it is unclear how well the results here will extrapolate to the other antigen tests due to variability in limit of detection or other test characteristics.

## Conclusion

Point-of-care SARS-CoV-2 antigen tests have the potential to significantly change the public health interventions needed to minimize the spread of COVID-19 by providing a better test to identify individuals that are likely to be shedding infectious virus and therefore transmit SARS-CoV-2. This will allow for rapid identification of asymptomatic COVID-19 cases and inform shorter periods of self-isolation for COVID-19 infected individuals. In addition, the low cost and scalability in low and middle-income countries associated with antigen-based testing will be an important tool in the diagnostic armamentarium to contain and suppress COVID-19 community transmission.

## Data Availability

Requests for data related to this study should be sent to Charles K. Cooper at Becton, Dickinson and Company.

## ACKNOWLEDGEMENTS

We thank the National Institute of Infectious Diseases, Japan, for providing VeroE6TMPRSS2 cells. We also thank Karen Eckert (Becton, Dickinson and Company, BD Life Sciences – Diagnostic Systems) for her input on the content of this manuscript and editorial assistance and Stanley Chao (Becton, Dickinson and Company, BD Life Sciences – Diagnostic Systems) for statistical support. The individuals acknowledged here have no additional funding or additional compensation to disclose. We are grateful to the study participants who allowed this work to be performed.

## AUTHOR CONTRIBUTIONS

All authors contributed to the interpretation of the data, critically revised the manuscript for important intellectual content, approved the final version to be published, and agree to be accountable for all aspects of the work.

## METHODS

### Study design and specimen collection

Prospective specimen collection, specimen use, and participant demographics for the parent study were described previously.^9^ This study involved the use of residual respiratory swab specimens from the previous antigen test Food and Drug Administration-Emergency Use Authorization (EUA) study, which occurred across 21 geographically diverse study sites, from June 5-11, 2020. Briefly, eligible participants were ≥18 years of age and had one or more self-reported COVID-19 symptoms between 0-7 days from symptom onset.^38,39^ Nasal swab specimens for use with antigen testing were collected only after the standard of care (SOC) swab. Nasopharyngeal (NP) swab specimens were collected after the nasal swab specimen for use with the RT-PCR assay (the laboratory reference standard in the EUA study); if an NP was collected as part of the SOC procedure at a collection site, the participant was given the choice of having an oropharyngeal (OP) swab specimen collected in lieu of a second NP swab for use with the RT-PCR assay. Overall, 76 specimen sets (consisting of one nasal and either one NP or one OP swab) were utilized from the original 251 evaluable specimen sets in the EUA study. The 76 specimens consisted of all 38 RT-PCR assay positive specimens, and 38, randomly selected RT-PCR assay negative specimens from the parent study. Specimens for the RT-PCR assay consisted of 71 NP swabs (37 and 34 positive and negative, respectively) and five OP swabs (1 and 4 positive and negative swabs, respectively). For the EUA study, reference testing was performed at TriCore Reference Laboratories while the antigen testing was performed internally at BD (San Diego, CA, USA). No study-related procedures were performed without an informed consent process or signature of a consent form. This research was performed in alignment with principles set forth by Good Clinical Practice guidelines and the Declaration of Helsinki. This article was prepared according to STARD guidelines for diagnostic accuracy studies reporting.^40^

### Test/assay procedures

#### Antigen test and RT-PCR assay

The antigen test (Becton, Dickinson and Company, BD Life Sciences—Integrated Diagnostic Solutions, San Diego, CA) and RT-PCR assay (Quidel Corporation. Athens, OH) were performed according to the manufacturers’ IFU.^19,41^ The only exception was that nasal swabs were shipped on dry ice (−70°) to the testing site prior to preparation for the antigen test. The RT-PCR assay reports cycle number in a manner that omits the first 10 cycles; here cycle numbers for the RT-PCR assay are reported with the addition of first 10 cycles.

#### SARS-CoV-2 virus culture

VeroE6TMPRSS2 was adapted from the VeroE6 cell line (ATCC CRL-1586) to express the TMPRSS2 protease at levels approximately 10-fold higher than that found in the human lung.^42^ The cells were cultured in complete medium (CM) consisting of Dulbecco’s modified Eagle Medium, supplemented with 10% fetal bovine serum (Thermo Fisher Scientific-Gibco, Waltham, MA), 1mM glutamine (Thermo Fisher Scientific-Invitrogen, Waltham, MA), 1mM sodium pyruvate (Thermo Fisher Scientific-Invitrogen, Waltham, MA), 100µg/mL penicillin (Thermo Fisher Scientific-Invitrogen, Waltham, MA) and 100 µg/mL streptomycin (Thermo Fisher Scientific-Invitrogen, Waltham, MA), at 37°C in a humidified chamber with 5% carbon dioxide. Cells were grown to 75% confluence in a 24 well plate format and the CM was removed and replaced with 150 µL of infection media (IM) which is identical to CM but with the fetal bovine serum reduced to 2.5%. One hundred microliters (100 µL) of the clinical specimen was added to each assay well and the cells were incubated at 37°C for two hours. The inoculum was then aspirated and replaced with 0.5 ml IM; the cells were then maintained at 37°C for four days. When a cytopathic effect was visible in most of the cells in a given well, the IM was harvested and stored at -70°C. The presence of SARS-CoV-2 was confirmed through quantitative RT-PCR as described previously,^7,43^ by extracting RNA from the cell culture supernatant using the Qiagen viral RNA isolation kit and performing RT-PCR using the N1 and N2 SARS-CoV-2 specific primers and probes in addition to primers and probes for human RNaseP gene using synthetic RNA target sequences to establish a standard curve.

### Probit models for probability of positive SARS-CoV-2 result

The RT-PCR assay was performed on serially diluted samples containing SARS-CoV-2 related genomic RNA prepared in universal transport media (containing human lung epithelial cells at 130,000 cells per mL) at concentrations ranging from 1.27 log10 copies/mL (cp/ml) to 4.27 log10 cp/ml (Table S1). The RT-PCR assay probability of positive result was fit using a probit model linking the Lyra results to viral RNA load. Linear regression was performed linking log10 cp/ml viral RNA load to Lyra Ct score using all samples with at least 3 log10 cp/ml (for which observed Lyra positivity was 100%).

Antigen test positivity and SARS-CoV-2 TMPRSS2 culture positivity (a surrogate for contagiousness), with RT-PCR confirmation, were fit with a probit model as a function of viral load, using results from the Veritor EUA study;^9^ RT-PCR assay Ct scores were used to estimate viral RNA loads, as described above. SARS-CoV-2 VeroE6 culture positivity linkage to viral load was inserted into the probit model for probability of a positive result using data from Huang et al (2020).^6^ Virus isolation in Huang et al was attempted for a total of 60 specimens, positive by RT-PCR for SARS-CoV-2. Of those, 23 were positive by culture. Ct scores of the SARS-CoV-2 envelope, nucleocapsid, and non-structural protein-12 RT-PCR targets were linked to the viral load (log10 cp/ml) through quadratic regressions. The empirical equation for envelope target Ct score was then used to calculate viral load for the 23 culture positive and 37 culture negative specimens (Figure S2). All analyses were performed using the R software system and the ggplot2 R package.^44,45^

**Table S1.** Corresponding values for Ct score and viral RNA load during limit of detection analysis involving Lyra SARS-CoV-2 RT-PCR assay-positive specimens

| **Viral Load**<br>**(copies/mL)** | **Log10**<br>**Viral Load** | **#Positive/**<br>**#Tested** | **PCR assay**<br>**Ct score; Mean (SD)** |
| --- | --- | --- | --- |
| 0 | - | 0/5 | - (-) |
| 19 | 1.27 | 0/9 | - (-) |
| 40 | 1.60 | 1/10 | 38.03 (-) |
| 86 | 1.94 | 1/9 | 38.83 (-) |
| 186 | 2.27 | 4/10 | 37.33 (1.20) |
| 400 | 2.60 | 8/10 | 35.89 (1.69) |
| 862 | 2.94 | 8/10 | 34.51 (1.62) |
| 1857 | 3.27 | 10/10 | 32.66 (1.07) |
| 4000 | 3.60 | 10/10 | 31.08 (0.75) |
| 8618 | 3.94 | 9/9 | 30.09 (1.15) |
| 18566 | 4.27 | 9/9 | 29.51 (1.45) |

**Figure S1.**
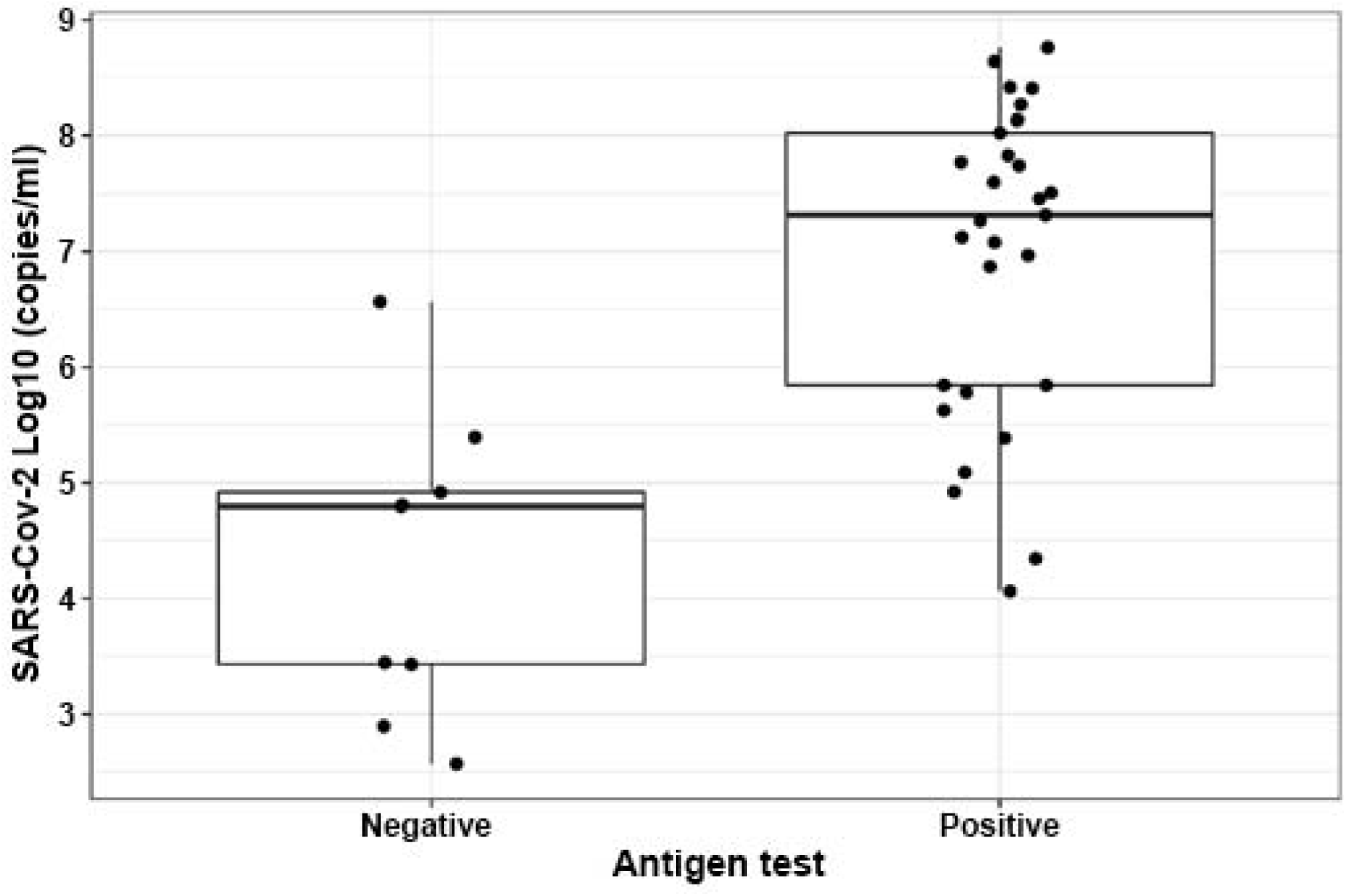
Box plots showing the median values for SARS-CoV-2 viral RNA loads from antigen-positive and -negative results within the 38 RT-PCR-positive results from the Veritor test EUA study. A two-sample t-test (2-tailed) analysis indicated a significantly higher mean (7.0 log10 cp/ml) for RT-PCR-positive/antigen-positive results compared to that (4.3 log10 cp/mL) for RT-PCR-positive/antigen negative results (p-value<0.001).

**Figure S2.**
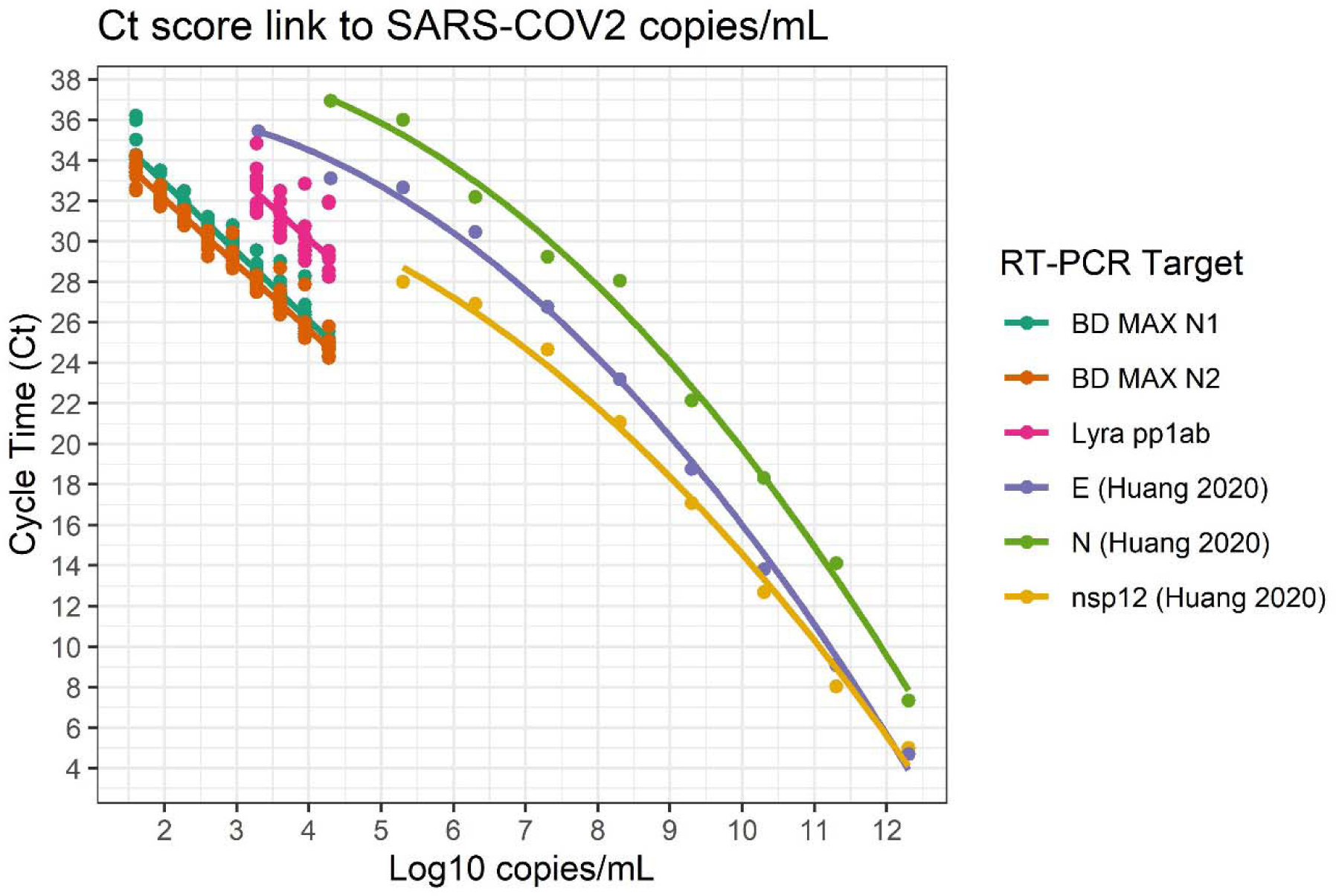
Relation of the RT-PCR Ct scores for the BD MAX assay, the RT-PCR assay, and the RT-PCR method used in Huang et al (2020)^6^ to viral load. Empirical equation for the RT-PCR assay Ct = 42.69 - 3.14 Log10 copies/mL. Empirical equation for the E target in Huang et al: Log10 copies/mL = 12.377 – 0.052 Ct – 0.005 Ct^2^

